# Modelling treatment response in tuberculosis early bactericidal activity trials

**DOI:** 10.64898/2026.01.19.26344385

**Authors:** Mairi CW McClean, Simon E Koele, Julia Dreisbach, Susanne Mirold-Mei, Fred Njeleka, Daniel Mapamba, Bariki Mtafya, Patrick PJ Phillips, Veronique R De Jager, Rodney Dawson, Kim Narunsky, Andreas H Diacon, Elin M Svensson, Norbert Heinrich, Francesco Paolo Casale, Michael Hoelscher

## Abstract

Culture-based monitoring of bacterial load is slow and susceptible to missing data, contributing to the length and cost of TB clinical trials. Non-culture-based alternatives, like the Tuberculosis Molecular Load Bacterial Assay (TB-MBLA), could represent a solution. Our objectives were to evaluate TB-MBLA as a biomarker in early bactericidal activity (EBA) studies and explore whether combining biomarkers with joint modelling could provide insight into underlying biological processes and reduce data loss. We generated TB-MBLA (LifeArc^(R)^) data from sputum samples from all 78 patients from the PanACEA BTZ-043 Phase Ib/IIa trial. In addition, we defined a joint biomarker as the first principal component derived from a probabilistic principal component analysis (pPCA) integrating TB-MBLA, colony forming units (CFU), and time-to-positivity (TTP) data. With TB-MBLA alone and the principal component 1 (PC1) marker we reevaluated the original stage IIa dose-response and stages Ib/IIa pharmacokinetics - pharmacodynamics (PK-PD) exposure-response analyses, applying linear and non-linear mixed models, respectively. For TB-MBLA, we could not detect an exposure-response effect in the PK-PD analysis, in contrast with CFU and TTP. When combining biomarkers, we observed a significant but less pronounced Emax exposure–response between days 0–3 compared with CFU and TTP alone. We also successfully applied pPCA as a modelling framework and show evidence that combining CFU and TTP in a joint latent component can improve detection of treatment effects compared with either biomarker alone.

**STUDY HIGHLIGHTS:** *What is the current knowledge on the topic?:* BTZ-043 is a first-in-class antimycobacterial compound with promising applications in drug-sensitive pulmonary tuberculosis. TB EBA trials are typically conducted using sputum culture assays for dose-exposure-response modelling, but the technical challenges of these assays limit their effectiveness.

*What question did this study address?:* How does the novel RT-qPCR assay TB-MBLA perform as a dose-exposure response marker for novel antitubercular compound BTZ-043 and what are the effects of joint modelling approaches in EBA modelling tasks?

*What does this study add to our knowledge?:* We did not observe a significant exposure response for BTZ-043 in TB-MBLA over the 14-day treatment window. Joint modelling with pPCA can overcome the issues of missing and contaminated data that are canonical of culture data from TB treatment monitoring cohorts.

*How might this change drug discovery, development, and/or therapeutics?:* Designing EBA trials should consider the drug-bacterial subpopulation axis of the specific study drug to maximise efficiency. Latent variable modelling techniques can be an effective and efficient framework for modelling *Mycobacterial tuberculosis* load in EBA trials.

## INTRODUCTION

A pivotal step in tuberculosis (TB) drug development is the early bactericidal activity (EBA) study, designed to evaluate the anti-mycobacterial effects of novel monotherapies or combination regimens over the first 7 or 14 days of treatment. At present, the biomarkers used to monitor treatment response are culture-based, colony forming units (CFU) on solid media and time-to-positivity (TTP) in liquid media. The technical complexity, susceptibility to contamination, data variability^1^ and long time to results^2^ have motivated the search for alternative treatment monitoring methods. In this context, the Tuberculosis Molecular Bacterial Load Assay (TB-MBLA), a real-time quantitative polymerase chain reaction (RT-qPCR)-based method, has emerged as a promising non-culture alternative for assessing treatment response. It has demonstrated applicability as a non-culture-based treatment monitoring assay^3^ and could overcome many limitations of culture-based biomarkers such as long incubation times and risk of contamination.

Each bactericidal assay captures distinct but overlapping bacterial subpopulations of *Mycobacterium tuberculosis* within the host^1,4,5^. These subpopulations differ in treatment response and growth dynamics^6,7^. CFU is a direct measure of the cultivable bacteria and TTP is an indirect time-to-event measure of dividing and aerobically metabolizing bacteria. In contrast, TB-MBLA quantifies the target 16s-rRNA, representing a quantification of all viable bacteria^8,9^; therefore, this assay could quantify additional non-cultivable populations otherwise missed by solid CFU and liquid TTP culture methods. While these biomarkers are often modelled separately, joint analysis may reveal shared dynamics of the overall bacterial population or highlight common treatment-response signals across assays^5^.

We generated TB-MBLA data from participants of the PanACEA-BTZ-043-02 phase Ib/IIa trial dose-ranging trial (clinical trial no NCT04044001), which enrolled 78 patients across 2 sites in South Africa, resulting in paired TB-MBLA, CFU and Mycobacterial Growth Indicator Tube (MGIT) TTP data for joint analysis. The primary results of the CFU and MGIT TTP data have been published elsewhere^10,11^. We then conducted a proof-of-concept analysis by applying a simple machine learning-based approach for joint modelling to the paired data, that is robust to missing data. The aims of this study were two-fold; to examine how TB-MBLA performs as marker of antimycobacterial drug efficacy; and to evaluate the effect of combining bactericidal assay data on the precision of model estimates.

## METHODS

### Patient recruitment and study design

All participants provided written informed consent, and the trial was approved by all required ethics committees (PharmaEthics: reference 190622615; Medical Faculty of LMU Munich: reference 19-0550; South African Health Products Regulatory Authority, reference 20190606).

The design and patient recruitment of the PanACEA-BTZ-043-02 phase Ib/IIa trial are described in detail in the main study publication^10^. Briefly, 24 and 54 patients were recruited in Phase Ib and IIa, respectively. In phase Ib, patients were assigned to ascending dose groups of BTZ-043 with daily doses of up to 1750 mg, with 3 patients per group and 6 patients in the highest dose group. In phase IIa, patients were randomized (3:3:3:2) to receive either 250, 500 or 1000 mg BTZ-043 daily or standard of care (HRZE).

### Bacterial load determination

During the trial, overnight sputum samples were obtained at Screening and Days 0, 2, 3, 4, 6, 8, 11 and 14 of treatment. Samples were homogenized and aliquoted into separate vials, each used once for a separate assay. As part of the original trial^10^, solid culture CFU counts were obtained from ten-fold dilutions of digested sputum inoculated on Middlebrook 7H11 selective agar plates (Media Mage, Johannesburg, South Africa). TTP was assessed from decontaminated sputum samples using the BACTEC^(TM)^ MGIT 960 system (Becton Dickinson, Johannesburg, South Africa). These assays were performed at the mycobacterial laboratory of TASK (South Africa). Assay data were log-transformed to approximate a normal distribution for downstream modelling tasks. Limits of quantification was set at 1.0 CFU/ml for solid CFU, and 600 hours (25 days) for MGIT TTP^12^.

For the present study, frozen stored sputum samples were processed using the LifeArc^(R)^ TB-MBLA PCR assay according to manufacturer guidelines^13^ at the National Institute for Medical Research-Mbeya Medical Research Center. PCR run-specific standard curves were generated to convert PCR quantification cycle (Cq) values to Starting Quantity (SQ) values, which are the number of copies of TB 16S per reaction (c/rxn). The SQ values represent the bacterial load and were log-transformed. The lower limit of detection was set at 128 copies per reaction (c/rxn).

### Probabilistic principal component analysis (pPCA)

To derive joint biomarkers aggregating information across assays, we used probabilistic principal component analysis (pPCA), a probabilistic variant of PCA formulated as a factor-analysis model (Equation 1), in which the data are assumed to arise from a lower-dimensional latent variable structure:

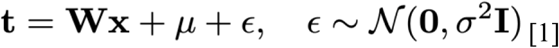

Here, **t** is the observed data, **x** the vector of latent variables, **W** the factor loading matrix, and **e** follows an isotropic Gaussian distribution, with **I** as the identity matrix. Traditional PCA seeks to reduce the dimensions of the original data into a subspace defined by the directions of highest variation using eigen decomposition of the sample covariance matrix. pPCA extends this by assuming a latent variable structure; here we estimate the conditional probability density of the observed data [Equation 2], and subsequently the latent variables [Equation 3].

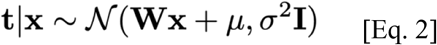

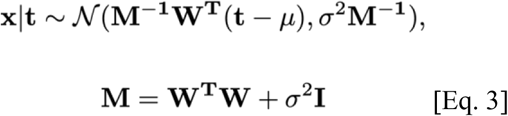

We obtain the principal components (PCs) loadings from the columns of **W**, and the PC scores per patient from **x**, through calculation of the mean of the posterior distribution [Equation 3] via Bayes theorem. Importantly, the probabilistic framework is applicable to datasets with missing data points ^14^, allowing inclusion of all patients with at least one available assay measurement at a given time point. Full details on the model and parameter estimation can be found elsewhere^14^. We applied the pPCA framework to compute the first principal component (PC1) from paired CFU, TTP, and TB-MBLA data, and then all combination pairs of CFU, TTP and TB-MBLA, using the pca-magic Python package^15^.

### Dose-exposure response modelling and statistical analysis

#### Dose-response

Descriptive dose response modelling was performed as in the original study^10^, using linear mixed effect methods, with random intercepts and random slopes in R (version 4.3.3). To assess the precision of model estimates, we calculated the coefficient of variation (CV) [Equation 4]:

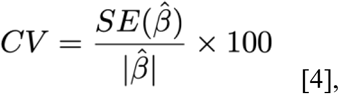

where 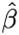 is the model coefficient and SE represents its standard error.

#### Exposure-response

For the PK-PD exposure-response analysis, the relationships between BTZ-043_total_ AUC_0-24_ and each response biomarker (TB-MBLA and PC1) were modelled using non-linear mixed effects methodology. BTZ-043_total_ represents the sum of BTZ-043 and its instable metabolite M2^16^, and the AUC values were derived from the population PK model developed as part of the original analysis^11^. In phase Ib, the AUC_0-24_ values were determined on Day 12, and in Phase IIa on Day 14.

The full PK-PD modelling process has been described in detail in the original study analysis^11^. Briefly, we examined linear and non-linear^17^ exposure-response relationships for the TB-MBLA and PC1 biomarkers and evaluated model fit by beginning with the simplest (linear) fit and testing successively complex exposure relationships via a likelihood ratio test (LRT) of the objective function value (OFV). Interindividual variability (IIV) was assumed to be log-normally distributed for the TB-MBLA data. The IIV for the intercept of PCA data was estimated on normal scale. The residual error model was additive on the log scale.

Stepwise covariate modelling was also performed as in the original analyses^11^, to examine the effect of baseline covariates on the exposure-response relationship. Age, weight, HIV status, and site were included.

As a qualitative measurement of statistical precision, we compared the magnitude of the decrease in OFV (dOFV) for the model selection process between the single and combined biomarkers. All modelling was performed using NONMEM 7.5 with Pirana 2.9.9 as graphical interface, and Perl-speaks-NONMEM 5.3.0 for additional functionalities. Visual predictive checks were employed to evaluate model fit. Model selection was performed using –3.84 dOFV as the threshold for significance for the LRT for model fit (corresponding to an alpha level of 0.05 [P<0.05]). Data management was performed using Python 3.10 in VSCode.

## RESULTS

We first fit the pPCA model to calculate PC1 term, and then compared this with the three individual biomarkers in the dose-exposure-response models from the original analysis.

### Bacteriological assays throughout treatment

Data from 78 participants across 8 timepoints were included. There were 0% contamination with the TB-MBLA assay, compared with 0.9% in MGIT TTP and 8.4% in solid CFU. All cases of missingness in the TB-MBLA were due to lack of sample availability. There were 2 TTP and 7 CFU samples above their respective limits of quantification, which were removed. There were no TB-MBLA datapoints above the limit of quantification. Tables of the summary statistics of all assays over time in each treatment group are included in the Supplementary materials (Tables S1 and S2). Figure 1 shows the per-patient trajectories for each bacteriological assay, including the combined marker, in each phase.

**Figure 1.**
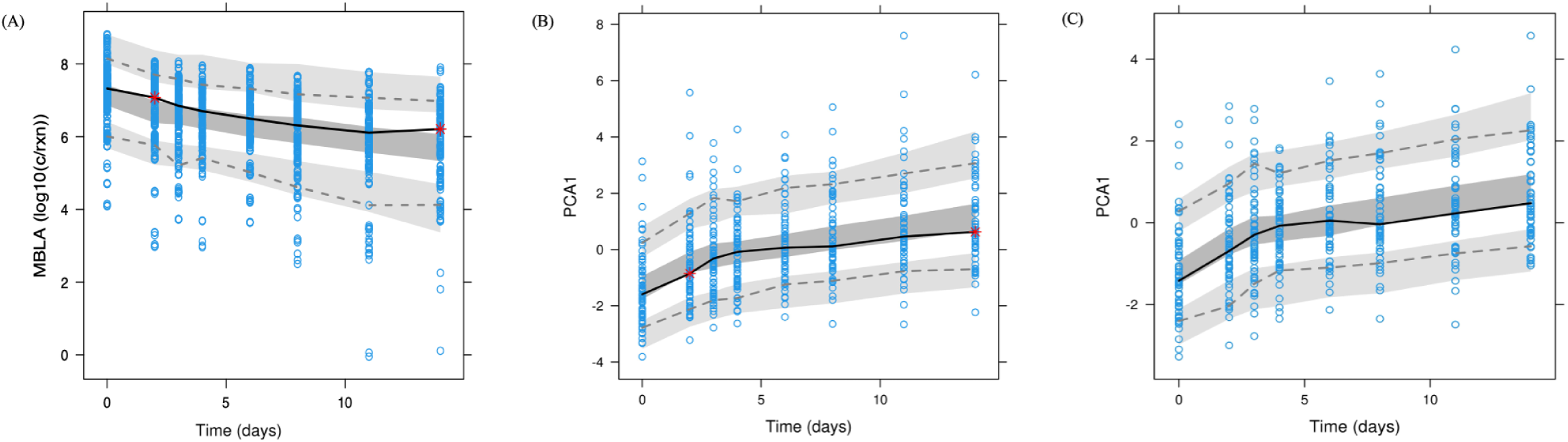
| Spaghetti plots of per-patient trajectories in each biomarker over time, by phase. The coloured lines represent the median over time. Breaks in individual lines represent missing data, either from sample availability or bacterial contamination.

### PC1 as a bacteriological biomarker

In our data, the first PC (PC1) explained a high proportion (86%) of the total variance of the data, was informed equally by each individual assay – as denoted by the similar magnitudes of loadings - and the direction of the loadings reflect the patterns observed in the original assays; the positive loading for MGIT TTP indicates that high MGIT TTP values correspond with high PC1 scores, and higher CFU and TB-MBLA assay values would result in lower PC1 values (Figure 2; Table 1). In context, lower PC1 values represent higher bacterial load, and higher PC1 values represent lower burden. As such, we hypothesized that PC1 represented a good summary metric of the joint distribution, with lower values indicating higher bacterial load. The second PC (PC2) represents the axis of second highest variance and is orthogonal to PC1; from the loadings, we observed that PC2 was primarily informed by TB-MBLA but did not explain a high amount of variance. Therefore, we considered only PC1 values for our downstream modelling tasks.

**Figure 2.**
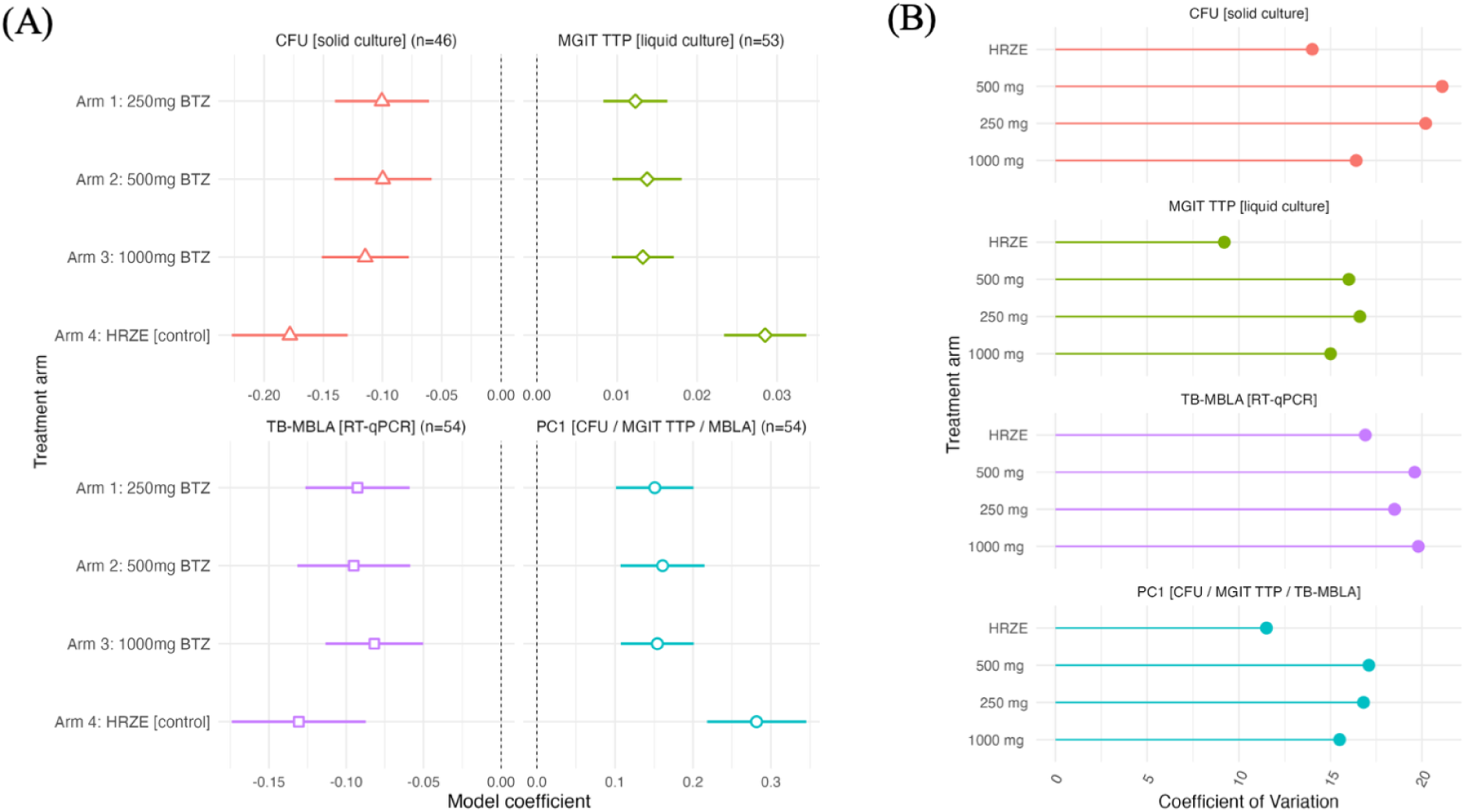
| Probabilistic Principal Component Analysis biplot. Each datapoint is from an individual patient at a single timepoint. The black lines - vectors - represent the relationship between the original variables on the first 2 PCs. Along the PC1 axis, the direction of the TB-MLBA and CFU vectors indicate that these biomarkers have negative loadings on this PC, and the MGIT TTP vector denotes a positive loading.

### Dose response models

We reproduced the dose-response modelling performed in the original study using the TB-MBLA and PC1 markers. Figure 3 reports the coefficients from the linear mixed model fixed effects, which represent the bactericidal slopes of each arm

**Figure 3.**
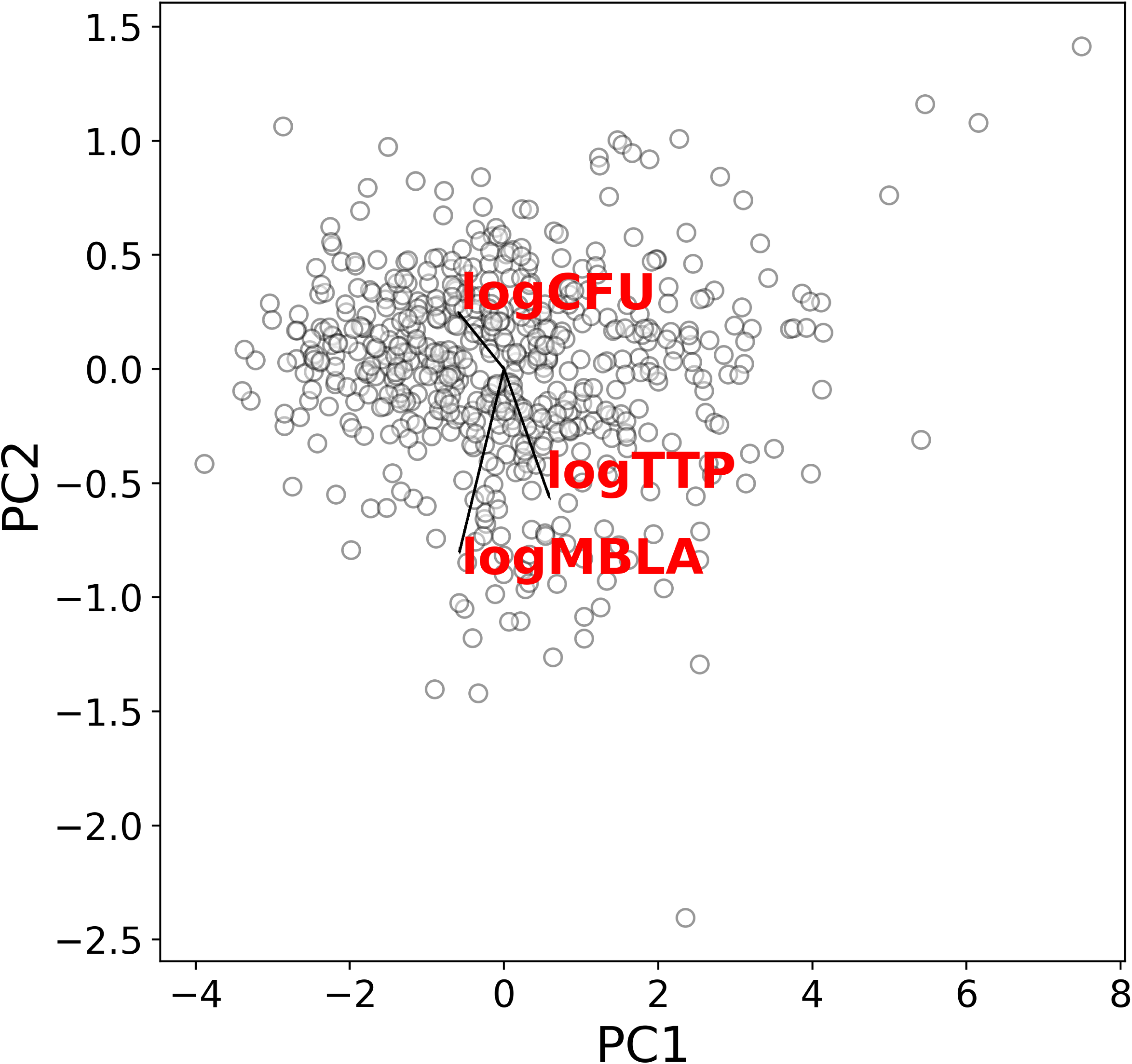
| Precision of linear mixed effect modelling by assay. Linear mixed models were constructed with a treatment-by-time interaction term as a fixed effect. (A) The resulting coefficients from the fixed effect represent the average slope per treatment arm. For MGIT TTP and PC biomarkers, a positive slope represents bactericidal activity, with greater values representing greater effect. Conversely, for CFU and TB-MBLA markers, a negative slope indicates presence of bactericidal activity, with greater values indicating greater effect. (B) CV values for each model coefficient. A smaller CV indicates greater precision of the model estimate. CFU, colony forming units, MGIT TTP, Mycobacterial Growth Indicator Tube Time to Positivity; TB-MBLA, Tuberculosis Mycobacterial Load Assay; CV, coefficient of variation.

As a measure of estimate precision, the CV for each treatment arm slope from each assay was calculated (Figure 3B; Table S3). TB-MBLA was comparable to solid CFU in terms of estimate precision, and MGIT TTP exhibited the greatest precision. The PC1 biomarker was more precise than the CFU and TB-MBLA single-assay models across all arms. The forest plots and CV values of the linear mixed models for all iterations of the pPCA algorithm (CFU + MGIT TPP; CFU + TB-MBLA, TB-MBLA + TTP) can be found in Figures S1 and 2.

### Exposure-response identified in PC1 marker, but not TB-MBLA

The TB-MBLA and PC1 markers were then examined in a PK-PD exposure-response framework. The complete model parameters for all models built are included in Supplementary Tables S4-S6.

#### TB-MBLA

The change in TB-MBLA log10 c/rxn over time was best described using bi-linear (stepwise linear) mixed model (LRT P<0.001) with a random intercept, suggesting a change in rate of bacterial decline over time. We estimated the node at 2 days, similar to what was found in the CFU and MGIT TTP models from the previous analysis^11^. Figure 4A shows a visual predictive check (VPC) indicating that the model fit the data well. We identified no significant exposure response of the BTZ-043_total_ AUC_0-24_ on either of the slopes, or over the entire 14-day treatment duration. We also assessed the relationship between TB-MBLA log10 c/rxn and BTZ-043 (parent); again, no exposure-response relationship could be identified. When examining the effects of baseline covariates on the exposure-response, weight was found to be associated with the second slope (post Day 2) in the bilinear model. No other association between any other parameter and baseline covariate was observed.

**Figure 4:**
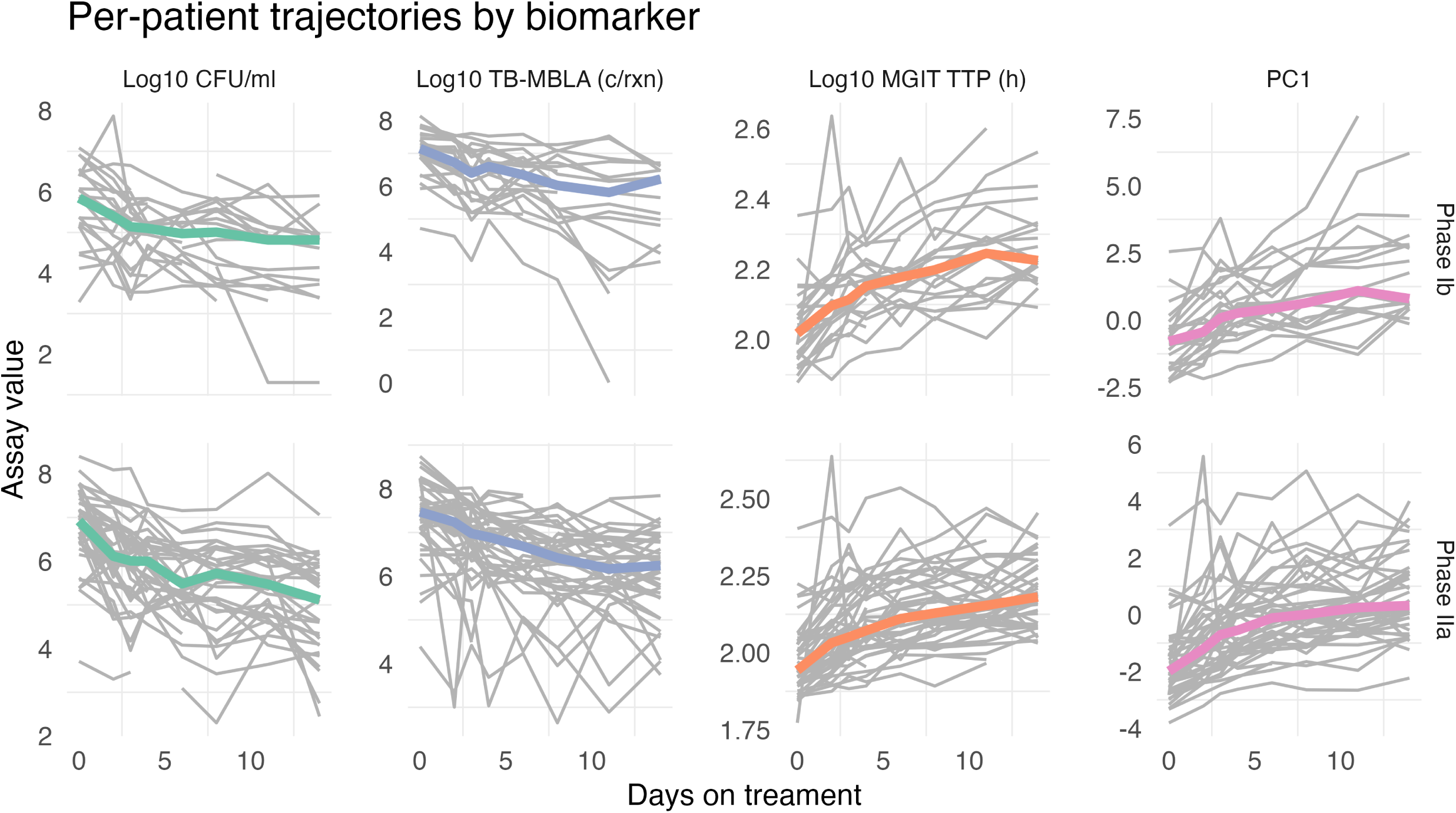
Visual predictive check plots of (A) TB-MBLA only, (B) 3-marker PCA (TTP + CFU + TB-MBLA) and (C) 2-marker PCA (TTP + CFU). Plots examining the model fit of each exposure-response model via simulations. Unbroken black line represents the median values per timepoint from the observed data, and the dashed lines the 5^th^ and 95^th^ percentiles of the observed data per timepoint. Grey areas represent the 95% confidence intervals for the 5^th^ and 95th percentiles, and the median, of the simulated data. Red points indicate where the observed median of the data falls outside of the range of predicted values.

#### pPCA biomarkers

We also applied the same model selection method to the PC1 values for each patient; a bi-linear model with the node at 3.2 days, with a significant exposure-response of BTZ-043_total_ AUC_0-24_ on the first slope, was selected as the best-fitting model (Fig. 4B), mirroring what was observed in the prior PK-PD analysis.

#### Exploring other biomarker combinations

Given the results from the TB-MBLA PK-PD model, we considered if the results observed in the PC1 model derived from all 3 assays may have been driven by signal in the CFU and MGIT TTP data. As such, we repeated the pPCA analysis using only CFU and MGIT TTP values and modeled the resulting PC1 as before. A bi-linear model with the node estimated at 2.8 days was selected as the best model, and significant exposure response with the BTZ-043_total_ AUC_0-24_ was identified on the first slope, until day 3 (Fig. 4C). Table 2 reports the results of our qualitative comparison of precision. A greater decrease in test statistic – here the objective function value (OFV) – indicates a greater effect size, translating to increased precision. The full table with all pPCA iterations is listed in Table S7.

When comparing all individual and combinations of biomarkers, the combination with the greatest ΔOFV was the CFU + MGIT TTP (-12.55), greater than both MGIT TTP and CFU alone. We also performed pPCA on all other possible combinations of biomarkers (CFU + TB-MBLA; MGIT + TB-MBLA). Addition of TB-MBLA resulted in a significantly smaller ΔOFV in all iterations, and there was no evidence to suggest an exposure-response in either the MGIT+MBLA or CFU+MBLA-derived PC1 markers, following the same model selection process outlined above.

## DISCUSSION

In the present study, we found that there was no observable exposure-response signal in TB-MBLA, in contrast with solid CFU and liquid MGIT TTP. We also demonstrated the utility of pPCA as a simple machine learning framework for modelling joint distributions that increased statistical resolution in PK-PD models compared with single markers.

Modelling approaches incorporating multiple biomarkers may allow us to simultaneously describe differential treatment effects in the different bacterial subpopulations present^5,18,19^. The PC1 from the pPCA model represents the direction of greatest variance of the dataset and a good summary metric of the overall distribution; it represents a holistic description of the overall bacterial population within the sample. We hypothesized that changes in PC1 may mirror a general treatment effect of a given compound, which was supported in part by our observation of a positive exposure-response signal in the PC1 data, and an increase in resolution of said signal compared with the single biomarkers. Additionally, the neat handling of missing data by pPCA should not be understated. The probabilistic nature allows for learning of the PCs even in patients with incomplete data, maximising downstream model estimate precision in a principled, data-driven manner.

In the PK-PD modelling in the present study, we did not observe an exposure response in the TB-MBLA data, which contrasted with the results from the previous PK-PD analysis in CFU and MGIT TTP alone^11^. Differences in signal by biomarker has been suggested to be treatment-induced^20^. BTZ-043 is a DprE1 enzyme inhibitor, inhibiting the synthesis of cell wall components^21,22^. Importantly, BTZ-043 has shown little efficacy against metabolically inactive cells^21^; cells which are theorized to comprise part of the population measured by TB-MBLA^3^. However, the lack of observable signal could also be due to increased noise within the assay. Future investigation of TB-MBLA data from longer studies with different compounds may help to investigate these hypotheses further and determine the full capacity of TB-MBLA to measure the long-term effects of antimycobacterial drugs.

The present study has several limitations: pPCA relies on gaussian distributional and temporal linearity assumptions, which may not reflect the true temporal covariance of the biomarkers ^23,24^. The impact may be limited in this case given the short timeframe of the study, but this might not hold in longer studies. These assumptions carry over to the PK-PD modelling task in which we assume the same exposure-response in each of the biomarkers. This is supported by evidence from the original description of the PK-PD response in each individual assay and has been observed for other drugs like rifampicin^11^. However, we concede that this assumption may prove inadequate when considering this method for other anti-mycobacterial compounds.

Despite the technical advantages of TB-MBLA over traditional culture methods, we did not observe an exposure-response signal for BTZ-043_total_ or BTZ-043, and these data did not add to the explanatory power of the combined biomarker models. However, we demonstrate that joint modelling via pPCA represents a practical approach to improve resolution of dose-exposure-response signal within an EBA study. Our pPCA-based approach increased the resolution of the exposure-response signal within an EBA study setting and proved robust to missing data, which is a common and limiting reality of culture-based treatment monitoring. Further work is required to develop this joint modelling framework to include more permissive inter-biomarker relationships, including non-linear relationships.

## Data Availability

Data collected for the study include individual participant data and a data dictionary defining each field in the set. PanACEA-TB will make de-identified data available on the TB PACTS trial repository. Researchers can apply to access these data on https://c-path.org/.

## ACKNOWLEDGEMENTS

We thank all the participants and site staff involved in the study and VdJ, RD, KN, and AD for patient recruitment at our trial sites. We also thank Katharina Limbeck and Jeremey Wayland for their statistical and mathematical support.

## AUTHOR CONTRIBUTIONS

MM and SK wrote manuscript, with all authors contributing to manuscript revisions. MH and FPC designed the research. FN, DM and BM performed the TB-MBLA assays. MM and SK performed the research and analysed the data, with support from ES and FPC.

## Code and data availability

The code to reproduce the pPCA model can be found in the study GitHub repository (https://github.com/mairi-mcclean/BTZ-043_pPCA.git).

## SUPPLEMENTARY INFORMATION TITLES

Table S1. Summary statistics by assay, by treatment arm for Phase Ib.

Table S2. Summary statistics by assay, by treatment arm for Phase IIa.

Table S3. CVs for the fixed effects from the dose-response mixed effect models.

Table S4. PK-PD model parameters for TB-MBLA.

Table S5. PK-PD model parameters for PC1 (MGIT TTP + CFU + TB-MBLA).

Table S6. PK-PD model parameters for PCA1 (MGIT TTP + CFU).

Table S7. Summary of exposure-response effects for BTZ-043 using different output markers.

Figure S1. Forest plot for fixed effect coefficients from linear mixed effect modelling for all iterations of PCA.

Figure S2. Coefficient of variation values for fixed effect coefficients from linear mixed effect modelling for all iterations of PCA.

